# Summaries, Analysis and Simulations of Recent COVID-19 Epidemic in Shanghai

**DOI:** 10.1101/2022.05.15.22273842

**Authors:** Lequan Min

## Abstract

**Background:** After successfully preventing the spread of five wave COVID-19 epidemics in Shanghai, Omicron and Delta variants have been causing a surge COVID-19 infection in this city recently. Summaries, analysis and simulations for this wave epidemic are important issues.

**Methods:** Using differential equations and real word data, this study modelings and simulates the recent COVID-19 epidemic in Shanghai, estimates transmission rates, recovery rates, and blocking rates to symptomatic and asymptomatic infections, and symptomatic (infected) individuals’ death rates. Visual simulations predict the outcomes of this wave Shanghai epidemic. It compares parallely with the recent mainland China COVID-19 epidemics (RMCE).

**Results:** The simulation results were in good agreement with the real word data at the end points of 11 investigated time-intervals. Visual simulation results showed that on the day 90, the number of the current symptomatic (infected) individuals may be between 852 and 7314, the number of the current asymptomatic (infected) individuals charged in the observations may be between 10066 and 50292, the number of the current cumulative recovered symptomatic infected individuals may be between 52070 and 74687, the number of the current cumulative asymptomatic individuals discharged from the medical observations may be between 63509 and 5164535. The number of the died symptomatic individuals may be between 801 and 1226.

- The transmission rate of the symptomatic infections caused by the symptomatic individuals was much lower than the corresponding average transmission rate of the RMCE.
- The transmission rate of the asymptomatic infections caused by the symptomatic individuals was much higher than the first 90 day’s average transmission rate of RMCE.
- The transmission rate of the symptomatic infections caused by the asymptomatic individuals was much lower than the first 60 day’s average transmission rate of RMCE, and was much higher than the last 60 day’s average transmission rate of RMCE.
- The transmission rate to the asymptomatic infections caused by the asymptomatic individuals was much higher than the corresponding average transmission rate of RMCE.
- The last 30 days’ average blocking rate to the symptomatic infections were lower than the last 30 days’ average blocking rates of RMCE
- The last 30 days’ average blocking rate to the asymptomatic infections were much higher than the last 30 days’ average blocking rate of RMCE. However the first 30 days’ average blocking rate to the asymptomatic infections were much lower than the first 30 days’ average blocking rate of RMCE.
- The first 37 days’ recovery rates of the symptomatic individuals were much lower than the corresponding first 70 days’ recovery rates of the symptomatic individuals of RMCE. The recovery rates between 38- and 52-days of the symptomatic individuals were much lower than the corresponding the recovery rates between 91- and 115-days of the symptomatic individuals of RMCE. The last week’s recovery rate was similar to the last week’s recovery rate of RMCE.
- The first 30 days’ average recovery rate recovery rate to the symptomatic individuals were much lower than the first 30 days’ average recovery rate recovery rate of RMCE. The last 30 days’ average recovery rate recovery rate of the symptomatic individuals were still much lower than the last 30 days’ average recovery rate of RMCE.

**Conclusions:** The last 30 days’ low blocking rates to the symptomatic infections, the first 30 day’s low blocking rates to the symptomatic infections to asymptomatic infections, the low recovery rates of the symptomatic and asymptomatic individuals, and the high transmission rate of the asymptomatic infections may be the reasons to cause the rapid spread of the recent Shanghai epidemic. It needs to implement more strict prevention and control strategies, rise the recovery rates of symptomatic and asymptomatic infections, and reduce the death rates for preventing the spread of this wave COVID-19 epidemic in Shanghai.

## 1 Introduction

Omicron and Delta variants have been causing surge COVID-19 infections in word wide. Many countries have experienced multiple outbreaks of the COVID-19 epidemics caused by the variants. As of 8 May 2022, over 514 million confirmed cases and over six million deaths have been reported globally [1].

After successfully preventing the spread of five wave COVID-19 epidemics in Shanghai, Omicron variants have broken through the COVID-19 prevention of this city recently. Analysis and simulations for this wave epidemic are important issues.

Since the outbreak of COVID-19 in Wuhan China, a large numbers of articles on modelings and predictions of COVID-19 epidemics have been published (for examples see [2–10]). Recently, the author has used several simple differential equation models to describe successfully the dynamics of spreads of the COVID-19 epidemics in mainland China ([11–14]).

Using the differential equation model and real word data, this study modelings and simulates the recent sixth wave COVID-19 epidemic (March 1 to April 30, 2022) in Shanghai, estimates transmission rates, recovery rates, and blocking rates to symptomatic and asymptomatic infections, and symptomatic individuals’ death rate. Visual simulations predict the outcomes of this wave Shanghai epidemic. It compares parallely with the recent mainland China COVID-19 epidemics (RMCE).

The rest of this paper is organized as follows. Section 2 introduces materials and methods. Section 3.1 summaries briefly the COVID-19 epidemic in Shanghai from January 4 to April 30, 2022. Section 3.2 modelings and simulates the dynamics of the Shanghai epidemic. Section 3.3 discuses the simulation results. Virtual simulations are implemented in Section 3.4. Concluding remarks are given in Section 4.

## 2 Materials and Methods

The dataset of the Shanghai COVID-19 epidemic from March 1, 2022 to April 30, 2022 was collected and edited from the Health Commission of Shanghai official website [15]. Using the differential equation model stimulates the outcomes of the numbers of the current symptomatic individuals, the current asymptomatic individuals charged in medical observations, the cumulative recovered symptomatic individuals, and the cumulative asymptomatic individuals discharged from medical observations, and the number of the cumulative died symptomatic (infected) individuals. The equation parameters were determined by so-called minimization error square criterion described in references [11, 14]. Using virtual simulations estimates the outcomes of the spreads of the recent COVID-19 epidemic in Shanghai. Simulations and figure drawings were implemented via Matlab programs.

## 3 Summaries, Analysis and Simulations of Recent Shanghai Epidemic

### 3.1 Summaries

- On January 4, 2022, the fifth wave Shanghai COVID-19 epidemic ended [12, 15]. The all symptomatic individuals in hospitals were recovered, except there were 8 asymptomatic individuals charged in medical observations.
- *From January 4 to January 12, 2022*. No new symptomatic infections were reported.
- *During the period from January 13 to February 22, 2022*. On January 13, two foreign input related symptomatic cases were reported. They were recovered on January 24 and January 25, respectively. On January 24 and January 26, two symptomatic individuals were reported. The first case may be infected by infected foreign import goods, and the second one was infected by the first individual. They were recovered on February 16 and February 22, respectively.
- *During the period from February 22 to February 28, 2022*. There were no reported symptomatic individuals; there were 10 asymptomatic individuals charged in medical observations.
- *During the period from March 1 to April 30, 2022* (sixth wave COVID-19 epidemic). The wave epidemic is still continuing. The outcomes of the numbers of the current symptomatic individuals (CSI) and the current asymptomatic individuals (CAI) charged in medical observations are shown in Fig. 1. The outcomes of the numbers of the cumulative recovered symptomatic individuals (CCSI), the cumulative asymptomatic individuals (CCAI) discharged from medical observations and the cumulative died symptomatic individuals are shown in Fig. 2. On March 1, one new symptomatic individuals was reported, there were ten cumulative asymptomatic individuals charged in medical observations, and there was one asymptomatic individuals discharged from the medical observations. On March 9 (day 8), one new asymptomatic individuals discharged from medical observations, there were 306 asymptomatic individuals charged in medical observations, and 23 current symptomatic individuals. The first symptomatic individuals’ recovery day was on March 19 (day 18), 10 asymptomatic individuals and 75 symptomatic individuals recovered on that day. There were 175 current symptomatic (hospitalized) individuals and 2099 asymptomatic individuals discharged from the medical observations. The asymptomatic infection turning point appeared on day 44 and 245812 asymptomatic individuals charged in medical observations. There were 12684 symptomatic individuals in hospitals. There were 2215 cumulative recovered symptomatic individuals. There were cumulative 44043 asymptomatic individuals discharged from the medical observations. The symptomatic infection turning point appeared on day 52 and 25010 symptomatic individuals in hospitals. There were 223682 asymptomatic individuals charged in medical observations. There were 223682 cumulative asymptomatic individuals charged in medical observations. There were 11965 cumulative recovered symptomatic individuals. There were 207287 cumulative asymptomatic individuals discharged from the medical observations. There were cumulative 48 died symptomatic individuals. The first died symptomatic individual appeared on day 47. On the investigated end day 60 (April 30) there were 20669 symptomatic individuals in hospitals and 162241 asymptomatic individuals charged in medical observations. There were 31888 cumulative recovered symptomatic individuals and 356338 cumulative asymptomatic individuals discharged from medical observations. There were 788 daily increased symptomatic individuals and 7084 daily increased asymptomatic individuals. There were 422 cumulative died symptomatic individuals..

**Figure 1.**
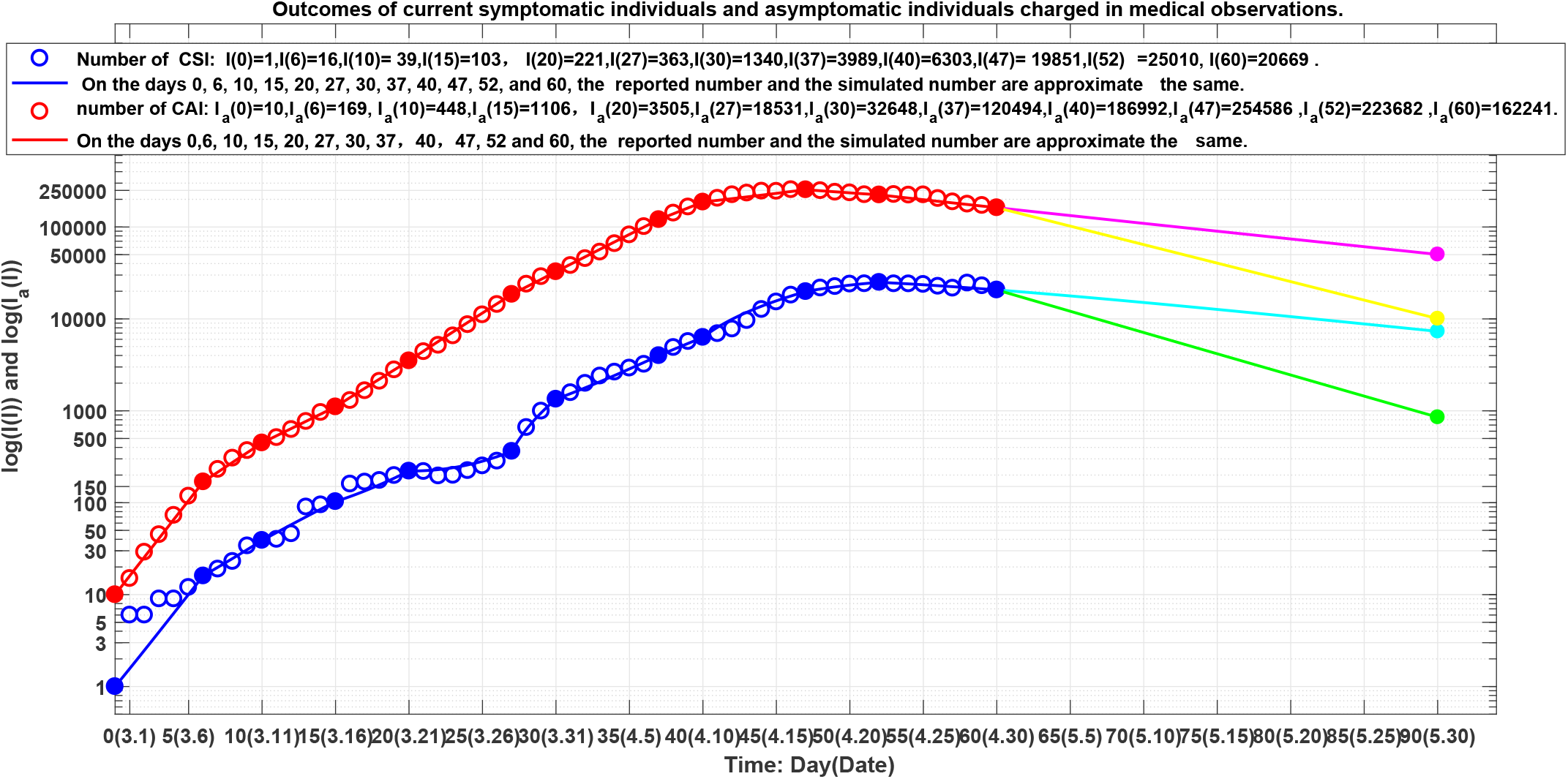
Blue circles: outcome of the number of the current symptomatic individuals (CSI), blue line: outcome of the corresponding simulation of equation (1). Red circles: outcome of the number of the current asymptomatic individuals (CAI) charged in medical medical observations, red line: outcome of the corresponding simulation of equation (1). The lines colored by cyan, magenta, green and yellow correspond the virtual simulation results of equation (1). See Section visual simulations for details.

**Figure 2.**
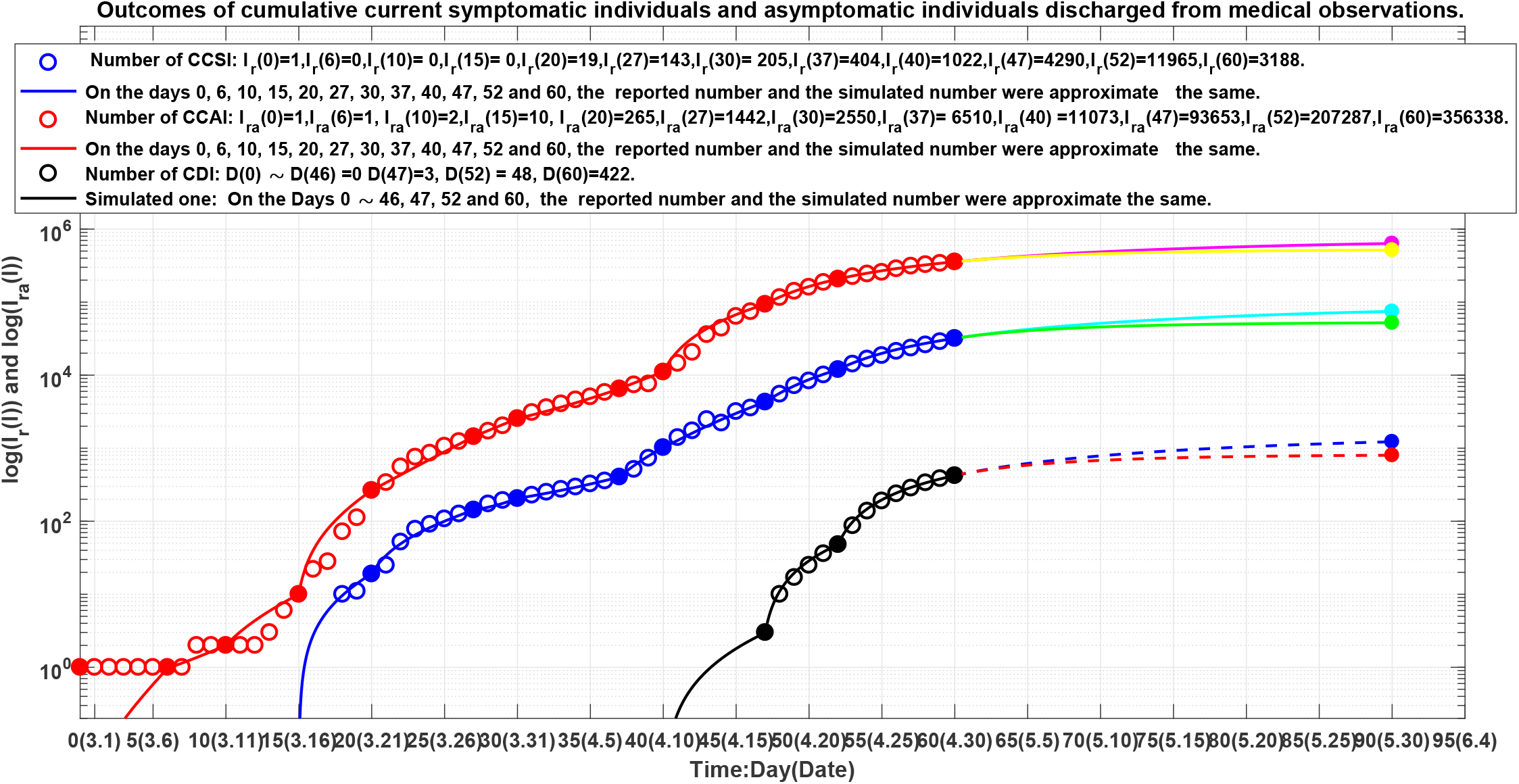
Blue circles: outcome of the number of the current cumulative recovered symptomatic individuals (CCSI), blue line: outcome of the corresponding simulations of equation (1). Red circles: outcome of the number of the current cumulative asymptomatic individuals (CCAI) discharged in medical medical observations, red line: outcome of the corresponding simulations of equation (1). Black circles: outcome of the number of the current cumulative died individuals (CCAI). Black line: outcome of the corresponding simulations of equation (1). The lines colored by cyan, magenta, green and yellow correspond to the virtual simulation results of equation (1). The dotted lines colored by blue and red corresponding to virtual simulation results for the cumulative died individuals. See Section Mainland Epidemic Virtual Simulations for details.

### 3.2 Simulations

In order to describe and understand the spread of an infectious disease, we need to set up a differential equation model to estimate the transmission rates and the blocking rates to symptomatic and asymptomatic infections. Assume that the process of the spread of an infectious disease are divided into *m* time intervals, representing different prevention control measures and treatment efficacy, respectively. Over the *lth* time interval, the model has the form (similar to [11, 13, 14]):

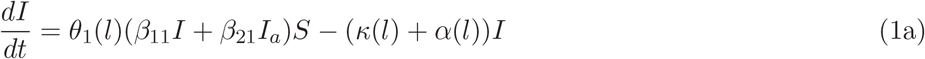

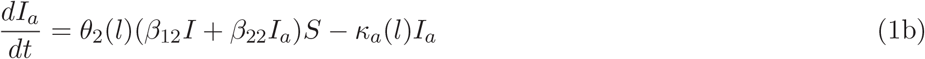

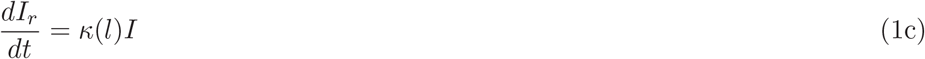

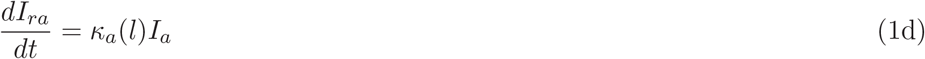

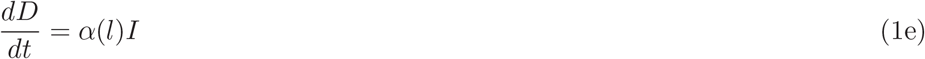

where Θ_1_(*i*) = (1 − *θ*_1_(*l*)) and Θ_2_(*l*) = (1 − *θ*_2_(*l*)) (*l* = 1, …, *m*) represent the blocking rates to symptomatic and asymptomatic infections, respectively. (*I*) and *I*_*a*_ represent the numbers of current symptomatic individuals and current asymptomatic individuals charged in medical observations, respectively. *I*_*r*_ and *I*_*ra*_ represent the numbers of current cumulative recovered symptomatic individuals and current cumulative asymptomatic individuals discharged from medical observations, respectively. *D* represents the number of current cumulative died symptomatic individuals. *β*_12_ and *β*_22_ represent the transmission rates of the symptomatic infections caused by symptomatic individuals and asymptomatic individuals, respectively. *β*_12_ and *β*_22_ represent the transmission rates of the asymptomatic infections caused by symptomatic individuals and the asymptomatic individuals, respectively. *S* represents susceptible population (can assume *S* = 1, see [11]). *κ*(*l*) and *κ*_*a*_(*l*) represent the recovery rates of the symptomatic individuals and the asymptomatic individuals, respectively. *α*(*l*) represents the death rate of the symptomatic individuals.

For Shanghai sixth COVID-19 epidemic, it can be assumed that the transmissions are divided into 11 time intervals (see solid points in Figs. 1 and 2). We need to determine the parameters of equation (1) for *l* = 1, 2, …, 11. Denote

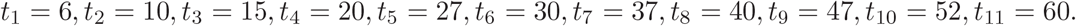

[*t*_*l*−1_, *t*_*l*_] is the *lth* time interval. Denote *I*_*c*_(*t*_*l*_) to be the number of the reported current symptomatic individuals, and *I*_*cr*_(*t*_*l*_) be the number of the reported current asymptomatic individuals charged in medical medical observations. Denote *I*_*cr*_(*t*_*l*_) to be the number of the reported current cumulative recovered symptomatic individuals, and *I*_*cra*_(*t*_*l*_) be the number of the reported current cumulative asymptomatic individuals discharged from medical medical observations.*D*_*c*_(*t*_*l*_) be the number of the reported current cumulative died individuals. Using the minimization error square criterion:

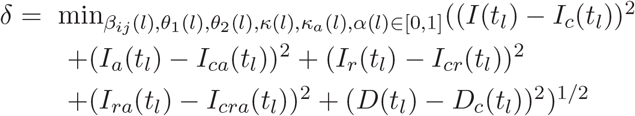

determines the *β*_*ij*_(*l*)^′^*s, κ*(*l*)^′^*s, κ*_*a*_(*l*)^′^*s*), *θ*_1_(*l*)^′^*s, θ*_2_(*l*)^′^*s*, and *α*(*l*)^′^*s*. The calculated parameters are shown in Table 1. The corresponding simulation results of equation (1) are shown in Fig. 1 and Fig. 2. Observe that the simulation results of equation (1) were in good agreement with the data of the COVID-19 epidemics. At the end points (see solid dots in Figs. 1 and 2) of the 11 investigated time-interval [*t*_*l*−1_, *t*_*l*_]^′^*s*, the simulated numbers and the actual reported numbers were approximate the same (Errors were less than one, respectively. See the solid blue lines, the red lines, and the black line in Fig. 1 and Fig. 2).

**Table 1.** Equation parameters of the COVID epidemics in Shanghai during 2022.3.1-2022.4.30. *β*_11_ = 0.035, *β*_12_ = 0.489, *β*_21_ = 0.0413, *β*_22_ = 0.4269.

| $l$ | Days | Dates | $\Theta_1(l)$ | $\Theta_2(l)$ | $\kappa(l)$ | $\kappa_a(l)$ | $\alpha(l)$ |
| --- | --- | --- | --- | --- | --- | --- | --- |
| 1 | 0-6 | 3.01-3.07 | 0% | 0% | 0 | 0.00288000 | 0 |
| 2 | 7-10 | 3.08-3.11 | 55.25% | 48.06% | 0 | 0.00090000 | 0 |
| 3 | 11-15 | 3.11-3.15 | 60.3 % | 61.18% | 0 | 0.00220000 | 0 |
| 4 | 16-20 | 3.16-3.20 | 70 % | 44.953% | 0.025470 | 0.02445000 | 0 |
| 5 | 21-27 | 3.21-3.27 | 90.11% | 42.18741% | 0.067400 | 0.01851900 | 0 |
| 6 | 28-30 | 3.28-3.30 | 67.218% | 53.961% | 0.025610 | 0.01483800 | 0 |
| 7 | 31-37 | 3.31-4.07 | 85.797% | 56.15122% | 0.011920 | 0.00840100 | 0 |
| 8 | 38-40 | 4.08-4.10 | 84.775% | 64.6835% | 0.040690 | 0.01005290 | 0 |
| 9 | 41-47 | 4.11-4.17 | 74.642% | 78.4521% | 0.036238 | 0.05400050 | 0.000033 |
| 10 | 48-52 | 4.18-4.22 | 75.807% | 85.3491% | 0.067660 | 0.09526660 | 0.000396 |
| 11 | 53-60 | 4.23-4.30 | 77.0718% | 88.2038% | 0.108624 | 0.09744147 | 0.002040 |

### 3.3 Results and Discussions

The equation parameters of the recent mainland COVID-19 epidemics (RMCE) are shown in Table 2 (see reference [16] amended version of reference [13]). From Table 1, Table 2, Fig. 1, and Fig. 2, it follows

**Table 2.** Equation parameters of the COVID epidemics in mainland China during 2021.12.31-2022.4.30 [16].

| $l$ | Days | $\beta_{11}(l)$ | $\beta_{21}(l)$ | $\beta_{12}(l)$ | $\beta_{22}(l)$ | $\Theta_1(l)$ | $\Theta_2(l)$ | $\kappa(l)$ | $\kappa_a(l)$ | $\alpha(l)$ |
| --- | --- | --- | --- | --- | --- | --- | --- | --- | --- | --- |
| 1 | 0-4 | 0.049056 | 0.072052 | 0.002102 | 0.094068 | 0% | 0% | 0.0172960 | 0.03107000 | 0 |
| 2 | 5-11 | 0.058000 | 0.072052 | 0.002102 | 0.094068 | 10.6% | 99% | 0.0452150 | 0.03131702 | 0 |
| 3 | 12-17 | 0.056600 | 0.072052 | 0.002102 | 0.094068 | 11.8 % | 99.6% | 0.0483000 | 0.00061700 | 0 |
| 4 | 18-30 | 0.044400 | 0.072052 | 0.004100 | 0.003990 | 49.59% | 40.95% | 0.0699400 | 0.01275000 | 0 |
| 5 | 31-43 | 0.077340 | 0.071000 | 0.002500 | 0.003000 | 51.02% | 72% | 0.0650000 | 0.05655000 | 0 |
| 6 | 44-48 | 0.124800 | 0.071000 | 0.025000 | 0.003000 | 51.15% | 71.6% | 0.0594400 | 0.04670000 | 0 |
| 7 | 49-55 | 0.124800 | 0.071000 | 0.020900 | 0.003000 | 25.04% | 72.377% | 0.0314600 | 0.03130000 | 0 |
| 8 | 56-64 | 0.124900 | 0.017800 | 0.059000 | 0.003000 | 31.976% | 70.29% | 0.0315600 | 0.01291000 | 0 |
| 9 | 65-70 | 0.125200 | 0.017800 | 0.141000 | 0.009000 | 26.02% | 21.07% | 0.0329400 | 0.00319000 | 0 |
| 10 | 71-77 | 0.287270 | 0.1 | 0.160680 | 0.009000 | 43.838% | 26.337% | 0.0240980 | 0.00468900 | 0.0000270 |
| 11 | 78-82 | 0.287270 | 0.1 | 0.160680 | 0.009000 | 73.263% | 37.524% | 0.0284910 | 0.01894300 | 0 |
| 12 | 83-90 | 0.287270 | 0.1 | 0.294230 | 0.009000 | 87.602% | 39.037% | 0.0398450 | 0.01839630 | 0 |
| 13 | 91-102 | 0.287200 | 0.1 | 0.500010 | 0.1 | 93.401% | 27.93% | 0.0867320 | 0.01816700 | 0 |
| 14 | 103-107 | 0.287200 | 0.1 | 0.5 | 0.1 | 90.252% | 45.973% | 0.0747380 | 0.06136523 | 0.0000240 |
| 15 | 108-110 | 0.287200 | 0.1 | 0.5 | 0.1 | 91.693% | 60.786% | 0.0711410 | 0.08755600 | 0.0002420 |
| 16 | 110-115 | 0.287270 | 0.1 | 0.5 | 0.1 | 93.337% | 56.63% | 0.0921265 | 0.08093320 | 0.0010987 |
| 17 | 116-120 | 0.287270 | 0.1 | 0.5 | 0.1 | 92.093% | 76.839% | 0.1199300 | 0.10006000 | 0.0017580 |

1. The transmission rate *β*_11_ of the symptomatic infections caused by the symptomatic individuals was much lower than the first 60 day’s average transmission rate *β*_11_ of RMCE (0.035: 0.082487), and was much lower than the last 60 day’s average transmission rate *β*_11_ of RMCE (0.0350: 0.26929).
2. The transmission rate *β*_12_ of the asymptomatic infections caused by the symptomatic individuals was much higher than the first 90 day’s average transmission rate of RMCE (0.489:0.0224), and was similar to the last 30 day’s average transmission rate of RMCE (0.4890:0.5000).
3. The transmission rate *β*_21_ of the symptomatic infections caused by the asymptomatic individuals was much lower than the first 60 day’s average transmission rate of the RMCE (0.0413:0.06488), and was much higher than the last 60 day’s average transmission rate of RMCE (0.0413:0.0909).
4. The transmission rate *β*_22_ of the asymptomatic infections caused by the asymptomatic individuals was much higher than the first 60 day’s corresponding average transmission rate of RMCE (0.4269: 0.0147), and was much higher than the last 60 day’s corresponding average transmission rate of RMCE (0.4269: 0.0596).
5. The last 30 days’ average blocking rate of Θ_1_(*l*)^′^*s* to the symptomatic infections were lower than the last 30 days’ average blocking rate of RMCE (79.5679:92.1553).
6. The last 30 days’ average blocking rate of Θ_2_(*l*)^′^*s* to the asymptomatic infections were much higher than the last 30 days’ average blocking rate of RMCE (74.5679:53.6328). However the first 30 days’ average blocking rate to the asymptomatic infections were much lower than the first 30 days’ average blocking rate of RMCE (41.7236:59.8875).
7. The first 37 days’ recovery rates *κ*(*l*)^′^*s* of the symptomatic individuals were much lower than the corresponding first 70 days’ recovery rates of the symptomatic individuals of RMCE. The recovery rates *κ*(*l*)^′^*s* between 38-and 52-days of the symptomatic individuals were much lower than the corresponding the recovery rates between 91- and 115-days of the symptomatic individuals of RMCE. The last week’s recovery rate was similar to the last week’s recovery rate of RMCE.
8. The first 30 days’ average recovery rate recovery rate of *κ*_*a*_(*l*)^′^*s* to the symptomatic individuals were much lower than the first 30 days’ average recovery rate of *κ*_*a*_(*l*)^′^*s* of RMCE (0.0106:0.0189). The last 30 days’ average recovery rate of *κ*_*a*_(*l*)^′^*s* of the symptomatic individuals were still much lower than the last 30 days’ average recovery rate recovery rate of RMCE (0.0530:0.0696).

### 3.4 Virtual simulations

Assume that after day 60 (April 30, 2022), it still keeps the the blocking rates Θ_1_(11) and Θ_2_(11), the recovery rates *κ*(11), *κ*_*a*_(11), and the death rate *α*(11) until day 90 (May 30, 2022). The simulation results of equation (1) are shown in Fig. 1 and Fig. 2 by cyan lines, magenta lines, dot blue line, respectively. Calculated results show that at day 90, the numbers of the current symptomatic and asymptomatic individuals reach about 7314 (see cyan solid dot in Fig. 1) and 50292 (see magenta dot in Fig. 1), respectively. The numbers of the cumulative recovered symptomatic individuals and cumulative asymptomatic individuals discharged in medical medical observations reach about 74687 (see cyan solid dot in Fig. 2), and 635059 (see magenta dot in Fig. 2), respectively. The number of death symptomatic individuals reach about 1226.

Furthermore assume that after day 60, it still keeps the recovery rates *κ*(11), *κ*_*a*_(11), and the death rate *α*(11) but increases the blocking rates (Θ_1_(*l*), Θ_2_(*l*)) ≡(99%, 99%) until day 90. The simulation results of equation (1) are shown in Fig. 1 and Fig. 2 by green lines yellow lines, and dot red line, respectively. Calculated results show that at day 90, the numbers of the current symptomatic and the asymptomatic individuals reduce about 852 (see green solid dot in Fig. 1) and 10066 (see yellow solid dot in Fig. 1), respectively. The numbers of the cumulative recovered symptomatic individuals and cumulative asymptomatic individuals discharged from medical observations reach about 52070 (see green solid dot in Fig. 2) and 516435 (see yellow solid dot in Fig. 2), respectively. The number of death symptomatic individuals reduce to about 801.

## 4 Concluding Remarks

The main contributions of this paper are summarized as follows:

1. It is the first time to summary and analyze the sixth Shanghai COVID-19 epidemic, and compare parallely with the recent mainland China COVID-19 epidemics (RMCE).
2. It uses model (1) to simulate the dynamics of the sixth Shanghai COVID-19 epidemic. The simulation results were approximate the same as the reported practical data at the end points of the investigated time intervals.
3. The simulation results suggest that:
  a. The transmission rate of the symptomatic infections caused by the symptomatic individuals was much lower than the corresponding average transmission rate of the RMCE.
  b. The transmission rate of the asymptomatic infections caused by the symptomatic individuals was much higher than the first 90 day’s average transmission rate of RMCE.
  c. The transmission rate of the symptomatic infections caused by the asymptomatic individuals was much lower than the first 60 day’s average transmission rate of RMCE, and was much higher than the last 60 day’s average transmission rate of RMCE.
  d. The transmission rate to the asymptomatic infections caused by the asymptomatic individuals was much higher than the corresponding average transmission rate of RMCE.
  e. The last 30 days’ average blocking rate to the symptomatic infections were lower than the last 30 days’ average blocking rates of RMCE
  f. The last 30 days’ average blocking rate to the asymptomatic infections were much higher than the last 30 days’ average blocking rate of RMCE. However the first 30 days’ average blocking rate to the asymptomatic infections were much lower than the first 30 days’ average blocking rate of RMCE.
  g. The first 37 days’ recovery rates of the symptomatic individuals were much lower than the corresponding first 70 days’ recovery rates of the symptomatic individuals of RMCE. The recovery rates between 38- and 52-days of the symptomatic individuals were much lower than the corresponding the recovery rates between 91- and 115-days of the symptomatic individuals of RMCE. The last week’s recovery rate was similar to the last week’s recovery rate of RMCE.
  h. The first 30 days’ average recovery rate recovery rate to the symptomatic individuals were much lower than the first 30 days’ average recovery rate recovery rate of RMCE. The last 30 days’ average recovery rate recovery rate of the symptomatic individuals were still much lower than the last 30 days’ average recovery rate of RMCE.
4. Virtual simulations suggest that
  - The evolution of the number of the current symptomatic individuals may be located in the region between the cyan line and the green line shown in Fig. 1. The evolutions of the number of the current asymptomatic individuals charged in medical observations may be located in the region between the magenta line and the yellow line shown in Fig. 1.
  - The evolution of the number of the current cumulative recovered symptomatic individuals may be located in the region between the cyan line and the green line shown in Fig. 2. The evolutions of the number of the current cumulative asymptomatic individuals discharged from medical observations may be located in the region between the magenta line and the yellow line shown in Fig. 2.
  - The evolutions of the number of the cumulative death symptomatic individuals may be located in the region between the blue dot line and the red dot line shown in Fig. 2.
  - On the investigated end day 90, the number of the current symptomatic individuals may be between 852 and 7314, the number of the current asymptomatic individuals charged in the medical observations may be between 10066 and 50292, the number of the current cumulative recovered symptomatic individuals may be between 52070 and 74687, the number of the current cumulative asymptomatic individuals discharged from the medical observations may be between 63509 and 5164535. The number of death symptomatic individuals may be between 1226 and 801.

In summary, it needs to implement more strict prevention and control strategies and rise the recovery rates of symptomatic and asymptomatic infections such that the outcomes of the numbers of the current symptomatic and asymptomatic individuals are near or below the green line and the yellow line shown in Fig. 1; the outcomes of the numbers of the current cumulative death symptomatic individuals are near or below the red dot line shown in Fig. 2.

It finds that different combinations of the nine parameters of Equation (1) may generate similar simulation results. Hence it needs further study to obtain better parameter combinations to interpret COVID-19 epidemics. It is not wise strategy to withdraw all prevention and control measures before the number of the all people have been cleared. 100% blocking rate to COVID-19 infection spread is key strategies for early clearance or reduction of epidemic spread [11–14].

An administration should at least maintain the prevention and control measures implemented one week after reaching the infection turning point of the numbers of the current hospitalized symptomatic individuals and the current asymptomatic individuals charged in medical observations [12–14].

## Data Availability

The dataset was collected and edited from the Health Commission of Shanghai official website， and National Health Commission of China. The 60 days' data
are needed to download from the links: https://wsjkw.sh.gov.cn/yqtb/index_2.html-https://wsjkw.sh.gov.cn/yqtb/index_7.html

https://wsjkw.sh.gov.cn/.

https://www.nhc.gov.cn/

https://wsjkw.sh.gov.cn/yqtb/index_2.html

https://wsjkw.sh.gov.cn/yqtb/index_3.html

https://wsjkw.sh.gov.cn/yqtb/index_4.html

https://wsjkw.sh.gov.cn/yqtb/index_5.html

https://wsjkw.sh.gov.cn/yqtb/index_6.html

## Data availability statement

Data are available on reasonable request. Please email the author.

## Ethical Statement

Not applicable/No human participants included.

